# Toward Accurate and Actionable Differential Diagnosis with Lean LLM Orchestration

**DOI:** 10.1101/2025.10.28.25338335

**Authors:** Ethan Yang

## Abstract

Large language models (LLMs) can assist clinicians with diagnostic reasoning, yet their autonomous diagnostic performance remains uncertain. We evaluated OpenMedicine AI, an LLM-powered diagnostic agent with a deterministic controller, on 302 New England Journal of Medicine Clinicopathological Conference (CPC) cases, a benchmark renowned for diagnostic difficulty. Models produced ranked differential-diagnosis lists. Accuracy was assessed by inclusion of the ground-truth diagnosis within the Top-n list (Top-n accuracy) and by Capture@K, an actionability metric that is “captured” if any of the Top-n differentials would appropriately lead a clinician to order the diagnostic test of record (DToR) or its immediate precursor.

Across 302 CPCs, OpenMedicine AI achieved 46.0% Top-1 and 79.1% Top-10 accuracy, outperforming AMIE (32.5%, 68.9%) and physicians (15.6%, 20.9%). Paired McNemar tests confirmed superiority at all thresholds (p < 10^-5^). For actionability, at Capture@10 it matched or exceeded AMIE in 97.0% of cases and physicians in 96.7%. It rescued 99 of 302 cases missed by physicians (odds ratio [OR] 16.5) and 44 missed by AMIE (OR 7.3), reducing misses by 31 and 13 per 100 cases, respectively. These gains correspond to a number needed to assess (NNA) of 3.21 versus physicians and 7.95 versus AMIE. A safety margin was evident already at Capture@3, with rescues outnumbering failures to rescue versus physicians (109 vs 15; OR 7.27; 95% CI, 4.24 to 12.47; p=8.7×10^-19^) and versus AMIE (61 vs 15; OR 4.07; 95% CI, 2.31 to 7.15; p=9.84×10^-8^), corresponding to 31 and 15 fewer misses per 100 cases, respectively.

These findings indicate that a lightweight, deterministic controller layered over state-of-the-art LLMs can narrow the gap between diagnostic recall and clinical actionability. By producing high-quality differentials and prioritizing rational next tests, this approach offers a scalable, resource-efficient path to improved diagnostic performance in high-complexity clinical scenarios.

## Introduction

Recent randomized trials show that while LLM assistance improves physician performance over conventional resources, LLM-alone (e.g. GPT-4) at times performs indistinguishably from physician+LLM on vignette tasks (−0.9%, 95% CI −9.0 to 7.2; p=0.80)^1,2^. In contrast, the autonomous medical LLM Articulate Medical Intelligence Explorer (AMIE) established an important reference for multi-specialty diagnosis by evaluating differential diagnosis (DDx) accuracies against known ground truth. Beyond clinician–LLM pairing, AMIE demonstrated higher Top-n (n>1) DDx accuracy than GPT-4 on a NEJM CPC subset^3^. Together, these results suggest that domain-optimized conversational systems such as Med-PaLM 2 and AMIE can meet or exceed general-purpose LLMs and clinician+LLM pairing^4^. Notably, although clinicians reported higher confidence when using the tool, clinician-assisted performance still lagged standalone AMIE, implying that DDx-only assistance may add cognitive load without proportional accuracy gain. Given that building high-quality DDx lists demands additional time and upskilling from clinicians, it is not obvious that DDx-only assistance is the most efficient path to real-world impact. Motivated by these observations, we take a pragmatic approach: we layer a low-overhead, deterministic pipeline on a frontier LLM and evaluate both (i) DDx list accuracy and (ii) capturability/actionability via Capture@K. We demonstrate superiority to physicians and state-of-the-art LLM baselines on both axes.

Related work by Hayat and colleagues introduced a proprietary multi-agent LLM that autonomously generated SOAP notes and reported strong diagnosis concordance in a virtual urgent-care setting^5^. However, the cohort was limited to US telehealth urgent care and skewed toward lower acuity presentations, and their own complexity subscore labeled about 3 percent as high. Clinicians reviewed the AI note before their visit, which is an acknowledged anchoring risk. The authors emphasized concordance rather than correctness, and there was no outcome level ground truth, so agreement with clinicians does not by itself establish clinical utility. By contrast, the NEJM Clinicopathological Conference (CPC) corpus we analyze concentrates diagnostically difficult cases; using the Hayat 0–10 complexity scale, 98% of this cohort falls within moderate-to-high complexity (scores 4–10). These differences motivate benchmarks that better capture genuine diagnostic difficulty while minimizing reliance on extensive human scaffolding.

Equally important, diagnostic testing is the next critical step in patient management, yet there is no widely accepted reference standard or benchmark to evaluate whether model-proposed tests are efficient and clinically appropriate. As described above, two common strategies for advancing AI in digital medicine are clinician augmentation with general-purpose LLMs and bespoke medical LLMs. Here we chart a third path by engineering a constraint-guided orchestration layer over general models. This approach raises diagnostic actionability without bespoke pretraining or workflow re-engineering.

To address both the accuracy and actionability gaps, we developed OpenMedicine AI, a deterministic, constraint-driven diagnostic agent that structures cases, proposes mechanism-based hypotheses, and reprioritizes them using evidence discriminators (organ distribution, imaging signatures, histology cues, and lab anchors). Chen and colleagues suggested potential clinical utility of generating further diagnostic tests, with helpfulness affirmed by an LLM grader and a blind physician panel^6^. We revisit 302 NEJM CPCs and extend evaluation beyond DDx accuracy to Capture@K, a novel metric that assesses whether the model’s Top-n differentials trigger the diagnostic test of record (DToR) or its immediate precursor, linking diagnostic reasoning to an actionable testing pathway.

## Results

As we hoped to evaluate the performance of OpenMedicine AI against other published models such as AMIE and GPT-4o, we compared the differentials produced by these models. To enable a like-for-like comparison, we first evaluated the 76 CPCs used in prior work^7^. The percentage of cases in which the correct final diagnosis appeared within the Top-n ranked differential diagnoses is shown for OpenMedicine AI (magenta), GPT-4o (blue), and AMIE (yellow). As expected, performance improved as n increased, with OpenMedicine AI consistently outperforming both comparators. At Top-1, OpenMedicine AI achieved 46.05% accuracy compared with 40.26% for GPT-4o and 26.32% for AMIE (Figure 1). At Top-10, OpenMedicine AI reached 81.58% accuracy versus 72.73% for GPT-4o and 57.89% for AMIE. On paired case comparisons across 76 CPCs, both AMIE and OpenMedicine AI were significantly more likely than clinicians to include the correct diagnosis in their Top-n lists (AMIE: Top-1 OR 3.50 [95% CI 1.10–14.60], p=0.0309; Top-10 OR 10.00 [3.11–51.21], p=1.40×10^−^□; OpenMedicine AI: Top-1 OR 9.33 [2.88–47.97], p=4.65×10^−^□; Top-10 OR 23.50 [6.15–199.73], p=4.36×10^−12^).

**Figure 1.**
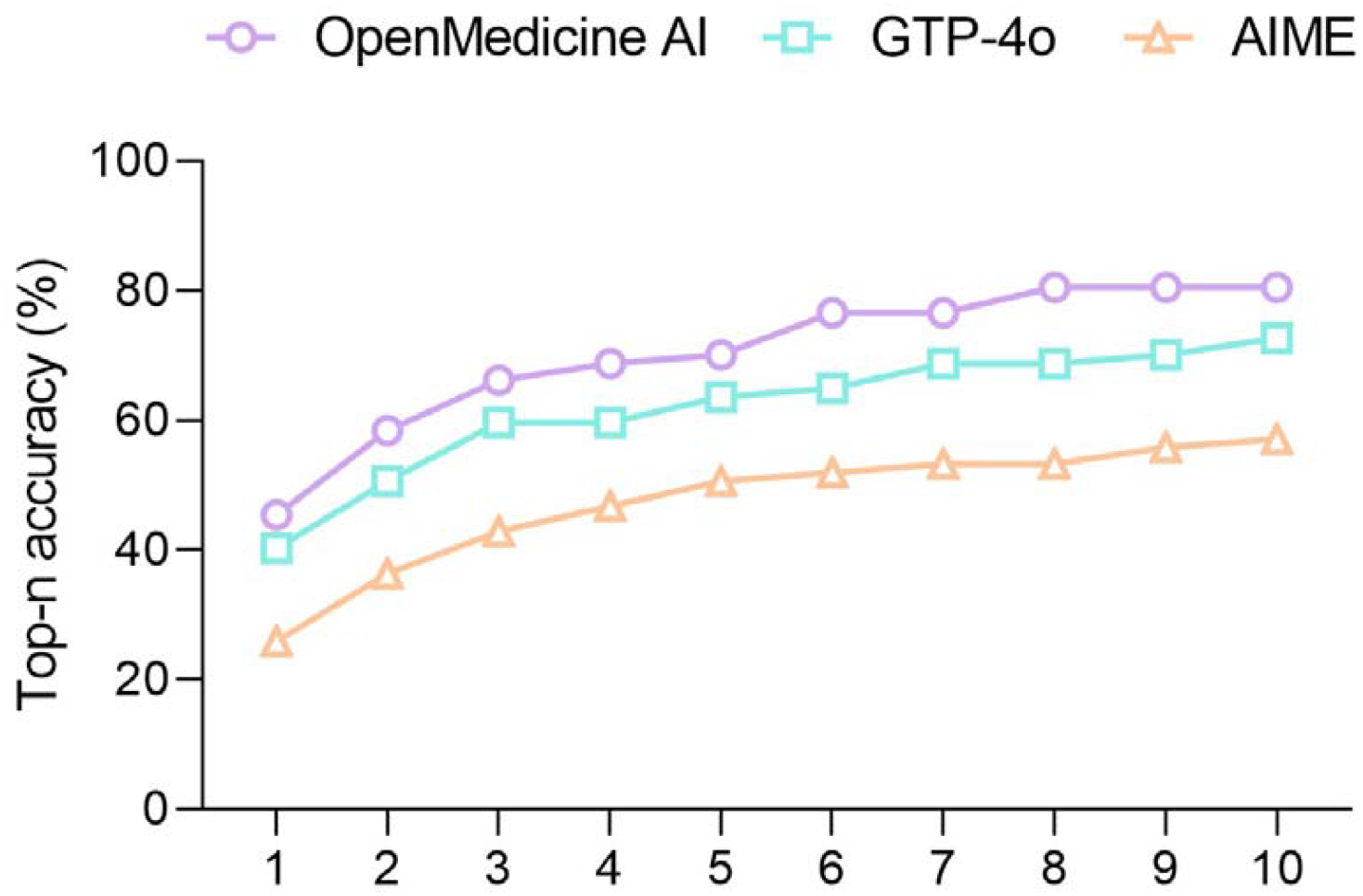
Top-n accuracy in DDx lists The percentage accuracy of DDx lists with the final diagnosis through automated evaluation by various LLMs (AMIE, GPT-4o, OpenMedicine AI)

**Figure 2.**
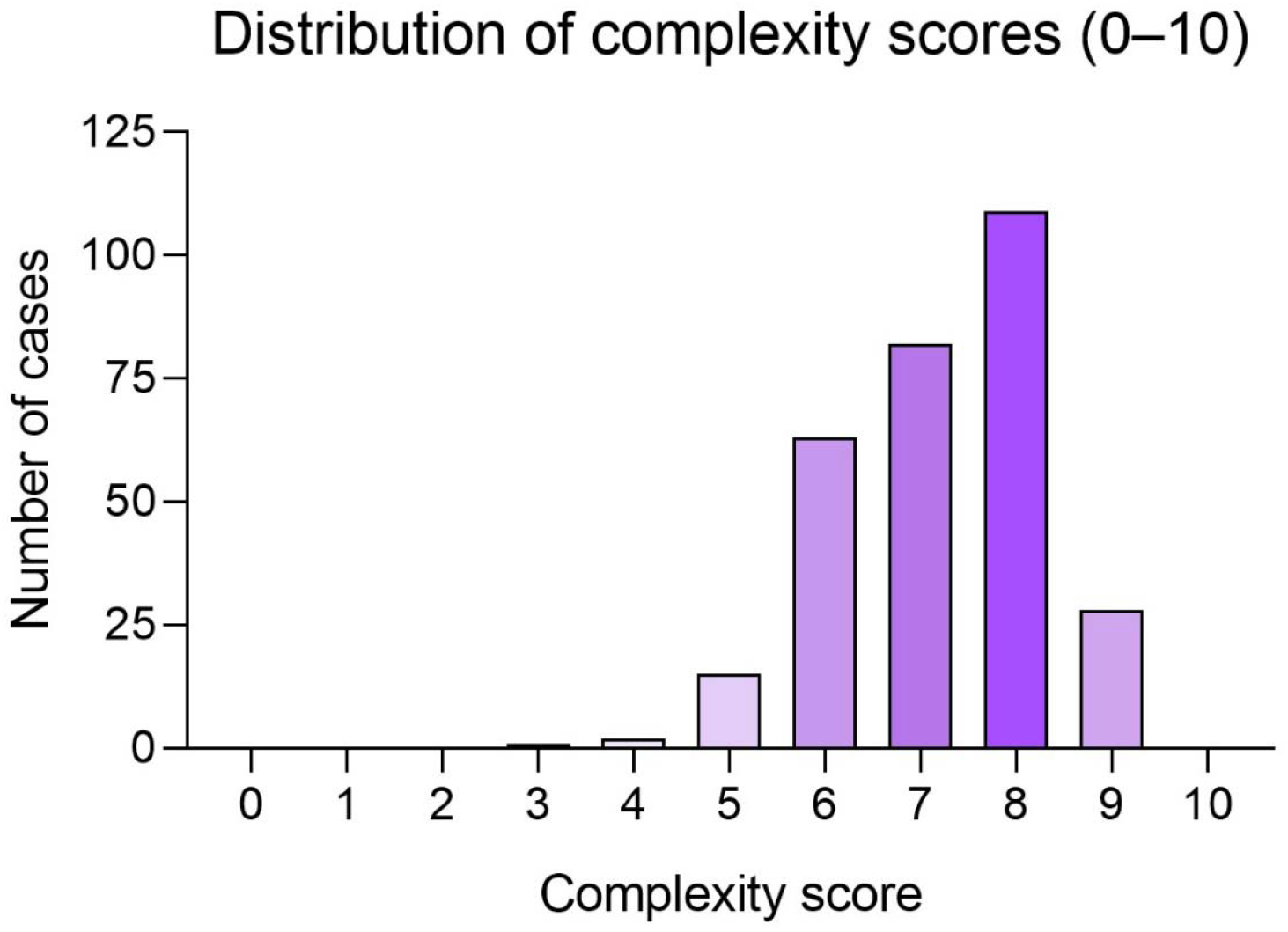
Distribution of case complexity scores (0-10) Among 302 cases, most scored 6–8 with a peak at 8, approximately 10% reached 9, and very few fell below 5, indicating an enrichment for higher-complexity presentations.

We then repeated the analysis across 302 CPCs and observed the same directional pattern, with OpenMedicine AI exceeding AMIE at each n (Top-1 46.03% and 32.45%; Top-10 79.14% and 68.87%; see Table 1). On paired case comparisons, OpenMedicine AI outperformed clinicians at Top-1 (McNemar exact p<2.2×10^−1^□; OR=7.57, 95% CI 4.34–13.22) and Top-10 (p<1×10^−1^□; OR=24.57, 95% CI 11.54– 52.32). OpenMedicine AI outperformed AMIE as well at Top-1 (McNemar exact p=2.03×10^−^□; OR=2.64, 95% CI 1.67–4.18) and Top-10 (p=3.13×10^−^□; OR= 2.45, 95% CI 1.50–4.03).

**Table 1.** Top-1 and Top-10 accuracy in DDx lists produced by AMIE and OpenMedicine AI.

| Model | Physicians |  | AMIE |  | OpenMedicine AI |  |
| --- | --- | --- | --- | --- | --- | --- |
|  | Top-1 | Top-10 | Top-1 | Top-10 | Top-1 | Top-10 |
| Full set (302 cases) | 15.56% | 20.86% | 32.45% | 68.87% | 46.03% | 79.14% |
| Partial set (76 cases) | 13.16% | 22.37% | 26.32% | 57.89% | 46.05% | 81.58% |
| Difference compared to full set | -2.40% | 1.51% | -6.14% | -10.98% | 0.02% | 2.44% |

Case complexity was assessed using the previously published 0–10 scale described by Hayat^5^. In this framework, scores reflect the inherent diagnostic difficulty, with common self-limiting conditions such as influenza rated at the lower end of the scale (0–3), and rare, multisystem, or diagnostically ambiguous conditions such as polymyositis rated at the higher end (7–10). To ensure comparability with prior work, we applied the same scoring rubric. The distribution of scores in our dataset was heavily weighted toward moderate-to-high complexity. Fewer than 5 cases scored below 5, while most cases clustered between 6 and 8, with a peak at 8. Approximately 10% of cases reached a score of 9, reflecting rare or diagnostically ambiguous and challenging presentations. Unlike the prior report where high-complexity cases comprised only a small minority, our dataset was enriched for diagnostically challenging presentations. These results highlight the superior diagnostic capacity of OpenMedicine AI consistently across a broad range of diagnostic depths.

To evaluate whether AI systems can identify the definitive diagnostic pathway rather than just list plausible conditions, we measured Capture@K, which quantifies the extent to which an agent’s Top-n differentials triggered the diagnostic test of record (DToR) or its immediate precursor. Scores are 1 when the DToR (or immediate precursor) is named, 0.5 for a pathway test that strongly advances toward the DToR, and 0 when there is no link to the diagnostic standard. In direct comparisons with physicians, OpenMedicine AI matched or outperformed Capture@3 in 93.05% of cases (281/302) (Table 2). Accuracy further increased with broader differential sets, reaching 94.70% (286/302) at Capture@5 and 96.69% (292/302) at Capture@10. Similar or higher rates were observed when compared to AMIE (93.71%, 95.03%, 97.02%; 95% CIs in Table 2). These findings demonstrate that OpenMedicine AI not only maintains high diagnostic precision at narrow Top-3 lists but also preserves performance robustness as the differential expands.

**Table 2.** Head-to-head win-or-tie rates for Capture@K.

| Metric | Comparison Target | OpenMedicine AI ≥ Target | Percentage of Cases | 95% CI |
| --- | --- | --- | --- | --- |
| Capture@3 | Physician | 281/302 | 93.05% | 89.57%, 95.64% |
| Capture@3 | AMIE | 283/302 | 93.71% | 90.35%, 96.17% |
| Capture@5 | Physician | 286/302 | 94.70% | 91.54%, 96.94% |
| Capture@5 | AMIE | 287/302 | 95.03% | 91.94%, 97.19% |
| Capture@10 | Physician | 292/302 | 96.69% | 93.99%, 98.40% |
| Capture@10 | AMIE | 293/302 | 97.02% | 94.42%, 98.63% |

For each case and $K \in \{3, 5, 10\}$ , win-or-tie ( ≥ ) counts cases where OpenMedicine Capture@K ≥ comparator. 95% CIs by Wilson method; $n = 302$ .

**Table 3.** Capture@K comparisons.

| Metric | Comparison Target | Probability of superiority | Wilcoxon statistic | 95% CI | p value (Wilcoxon, two-sided) |
| --- | --- | --- | --- | --- | --- |
| Capture@3 | Physician | 0.6523 | 7888 | [0.6192, 0.6854] | 2.03e-15 |
| Capture@3 | AMIE | 0.5745 | 2761 | [0.5464, 0.6026] | 8.82e-07 |
| Capture@5 | Physician | 0.6507 | 6830 | [0.6192, 0.6821] | 6.88e-16 |
| Capture@5 | AMIE | 0.5762 | 2269 | [0.5513, 0.6026] | 8.33e-06 |
| Capture@10 | Physician | 0.6523 | 5894 | [0.6242, 0.6821] | 8.59e-17 |
| Capture@10 | AMIE | 0.5646 | 1401 | [0.5414, 0.5877] | 1.06e-06 |

At Capture@3, the score distribution further illustrates these differences (Figure 3A). OpenMedicine AI produced the highest proportion of direct matches (score = 1) and the fewest incorrect captures (score = 0). Physicians more often failed to identify the diagnostic test of record, while AMIE showed a greater tendency toward partial pathway captures (score = 0.5). Similar patterns were recapitulated at Capture@5 and Capture@10, where the margin of superiority widened further (Figure 3B and 3C). Case-level probability of superiority (Capture@K): OpenMedicine AI vs baselines

**Figure 3.**
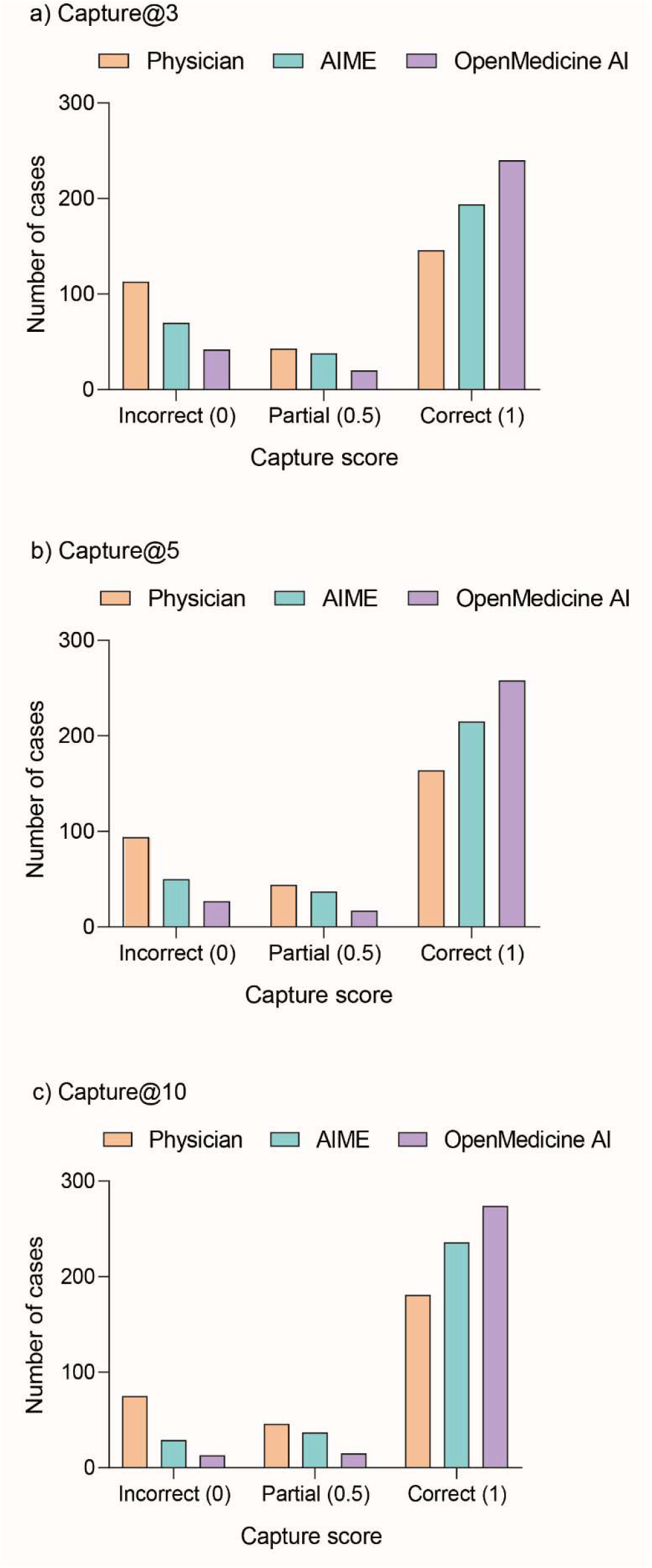
Distribution of capture scores at Top-n (n = 3, 5, 10; n=302) for Physicians, AMIE, and OpenMedicine AI Each differential diagnosis was mapped to the diagnostic test it would trigger and scored against the Diagnostic Test of Record (DToR). Scores were defined as follows: 1 = the DToR or its immediate precursor test was captured; 0.5 = a test was proposed that strongly suggested the pathway to the DToR but was not the immediate gateway; 0 = no meaningful link to the DToR.

**Figure 4.**
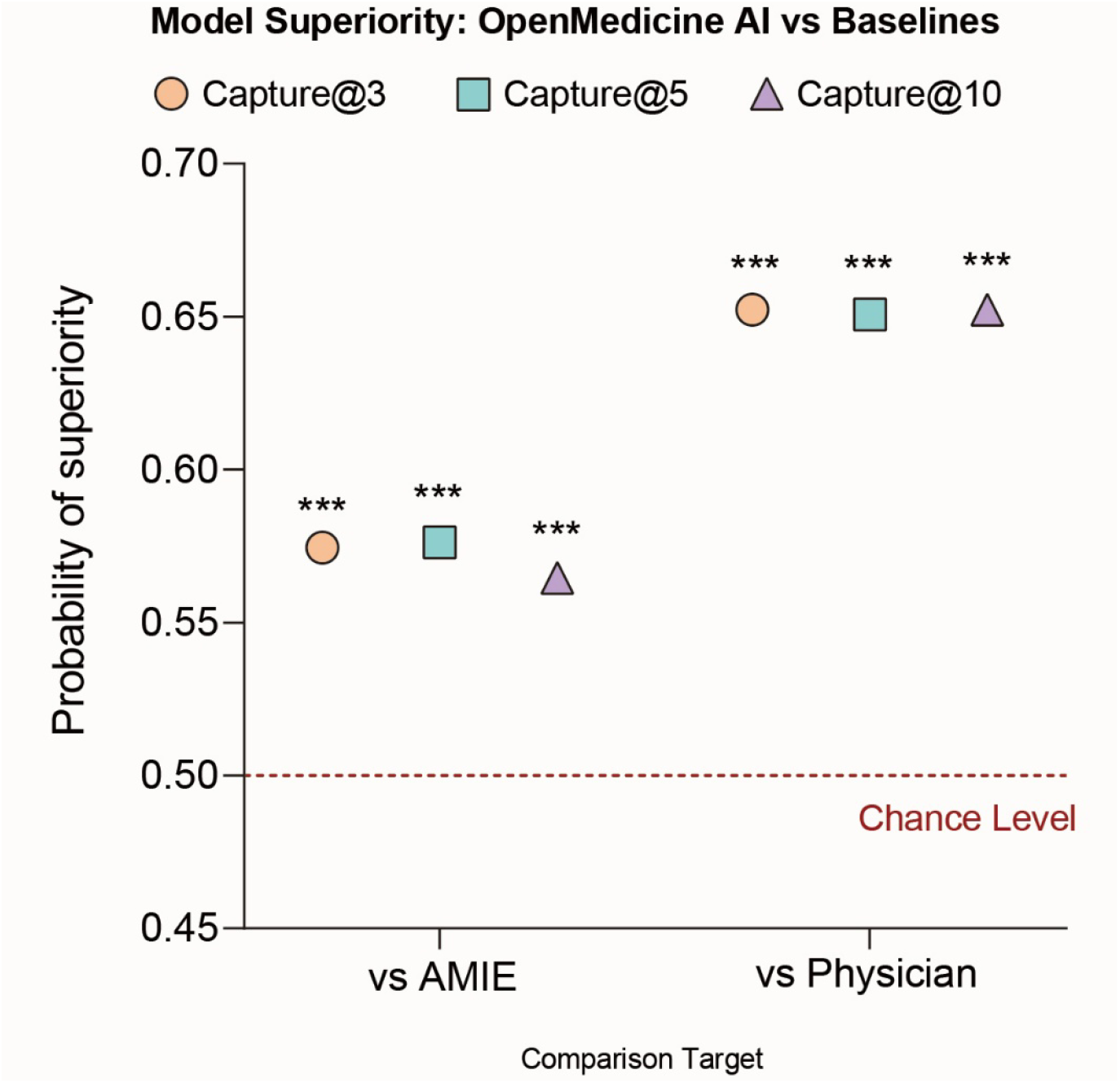
Case-level superiority on Capture@K Probability of Superiority (PS), Wilcoxon signed-rank statistic, 95% CI, and two-sided p value; PS>0.50 favors OpenMedicine AI (0.50=chance). ^***^ indicates p<0.001.

Each differential diagnosis was mapped to the diagnostic test it would trigger and scored against the Diagnostic Test of Record (DToR). Scores were defined as follows: 1 = the DToR or its immediate precursor test was captured; 0.5 = a test was proposed that strongly suggested the pathway to the DToR but was not the immediate precursor; 0 = no meaningful link to the DToR.

Across all three K thresholds, OpenMedicine AI outperformed with robust performance against both physicians and AMIE on case-level Capture. Paired Wilcoxon signed-rank tests (two-sided) compared OpenMedicine AI with each comparator at each K. We report the probability of superiority, the common-language effect size, interpreted as the probability that OpenMedicine AI exceeds the comparator among non-tied pairs using the Wilcoxon statistic, and the two-sided p value. Against AMIE, OpenMedicine AI showed consistent advantages at Capture@3 with 57.5%(p=8.82×10^−^□), Capture@5 with 57.6%(p=8.33×10^−^□), and Capture@10 with 56.5%(p=1.06×10^−^□)

To pinpoint where the systems diverged, we analyzed discordant pairs where one method was correct, and the other was not. A rescue when OpenMedicine AI was correct and the comparator was not and a failure to rescue for the reverse. Under the strict scoring rule (scores 0 or 0.5 counted as miss; n =302; ties excluded), rescues substantially outnumbered failure to rescues with either comparison. Against physicians, rescues: failure to rescues were 109:15 at Capture@3 (matched OR 7.27; 95% CI 4.24–12.49) and 99:6 at Capture@10 (OR 16.50; 95% CI 7.23–37.60), (Figure 5). Against AMIE, the corresponding counts were 61:15 at Capture@3 (OR 4.07; 95% CI 2.31–7.15) and 44:6 at Capture@10 (OR 7.33; 95% CI 3.13–17.20); all two-sided McNemar P < .001.

**Figure 5.**
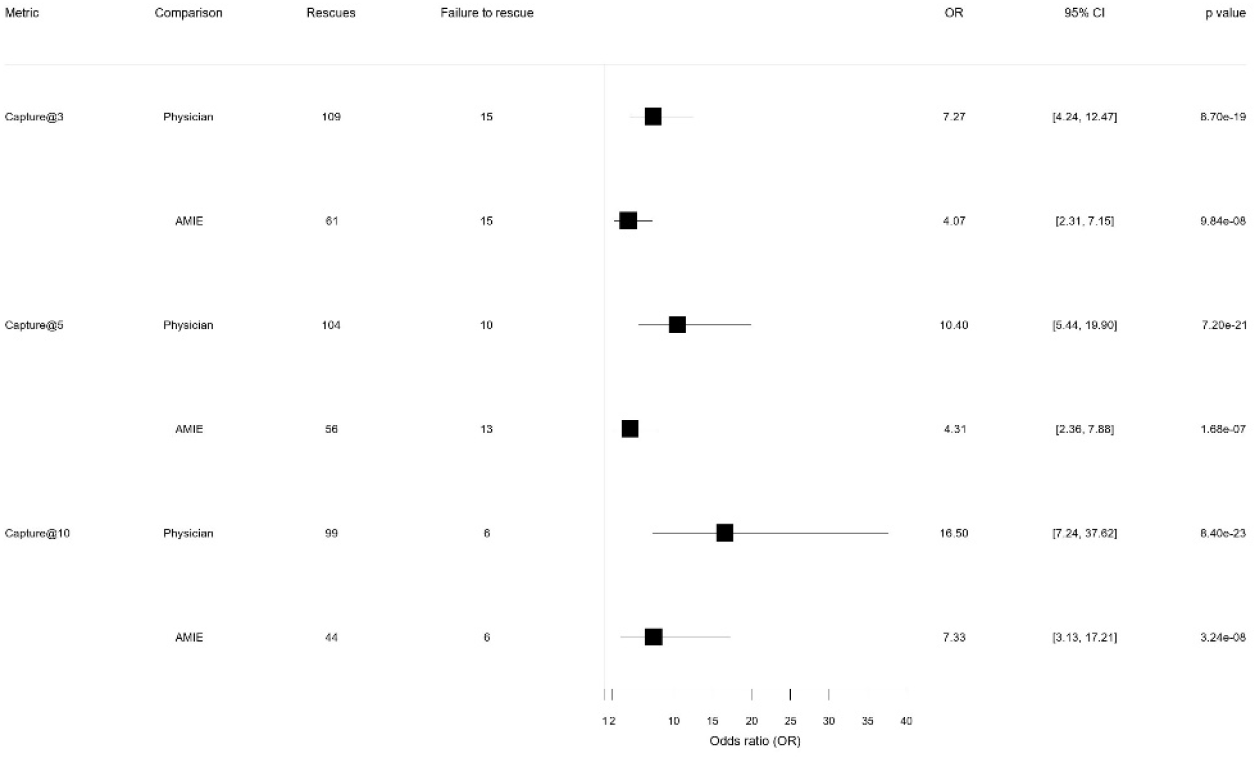
Discordant-pair analyses at Capture@3, Capture@5 and Capture@10 (n = 302) We report a rescue when OpenMedicine AI was correct and the comparator was not, and a failure to rescue for the reverse; We report matched OR (wins/losses), 95% CI, and two-sided McNemar p value.

At Capture@3, catastrophic misses fell by 109 cases relative to physicians (31.1 per 100, Figure 5), corresponding to a number-needed-to-assess (NNA) of 3.21 (diagnostic analog of NNT). This advantage was recapitulated at Capture@10, with 99/302 cases prevented (30.8 per 100; NNA= 3.25). Against AMIE, reductions were 46 cases at Capture@3 (15.2 per 100; NNA= 6.57) and 38/302 at Capture@10 (12.6 per 100; NNA= 7.95). Absolute lift at Top-10 (paired risk difference) was +30.8 percentage points vs physicians (95% CI +25.5 to +36.8) and +12.6 points vs AMIE (95% CI +8.3 to +16.9).

## Discussion

Clinician-facing evidence assistants such as AMIE and OpenEvidence are rapidly proliferating; however, randomized vignette data indicate that clinician+LLM assistance does not necessarily outperform a standalone LLM, suggesting that DDx-only support may tax cognitive load without net accuracy gains unless coupled to action-oriented pipelines^3^. These two trajectories are accelerating in parallel: bespoke, domain-optimized assistants and rapidly advancing generalist models. It is not obvious which yields superior clinical utility.

We developed OpenMedicine AI, a novel LLM-derivative optimized for clinical reasoning and diagnosis. Compared with AMIE, OpenMedicine AI generated more accurate differential-diagnosis (DDx) lists, producing outputs that were both more appropriate and more likely to include the correct final diagnosis. This advantage was even more pronounced when benchmarked against physician-generated differentials.

The NEJM CPCs used in this study are widely recognized for their diagnostic difficulty and have long served as a standard for evaluating clinical reasoning. Using the Hayat complexity scale, this dataset was substantially more challenging than prior benchmark sets, which were weighted toward simpler presentations. Within this enriched cohort, OpenMedicine AI consistently outperformed both AMIE and unassisted board-certified physicians across Top-1 and Top-n accuracy. While superior Top-1 and Top-10 performance underscores OpenMedicine AI’s strength in constructing DDx lists, the results also suggest diminishing returns at higher ranks, a plausible ceiling effect given the limited information available in many CPC vignettes.

Clinician feedback in the Nature publication suggested that AMIE as assistance was “most helpful for simpler cases with specific keywords or pathognomonic signs,” yet it struggled to integrate findings holistically. In that setting, clinicians improved results by filtering AMIE’s outputs through their own reasoning and judgment. However, even with this assistance, their performance still fell short of the AMIE model operating alone. This paradox underscores the limitations of relying on post-hoc clinician filtering of weaker LLMs and upskilling demands on clinicians without a commensurate gain in accuracy. Our findings with OpenMedicine AI suggest that the more effective path forward is to develop models that natively incorporate structured clinical reasoning, rather than depending on clinicians to compensate for gaps in model performance.

Because CPCs function as diagnostic stress tests rather than mirrors of everyday workflow, we extended our evaluation beyond diagnosis to Capture@K, a measure of whether an agent’s reasoning leads toward the diagnostic test of record (DToR). Across all thresholds, OpenMedicine AI more frequently produced differentials that pointed directly to the DToR or its immediate precursor, surpassing both physicians and AMIE. These findings show that OpenMedicine AI is not only proficient at generating accurate diagnoses but also supports clinically actionable reasoning pathways that guide testing and decision-making. Earlier work showed moderate helpfulness from multi-agent conversational LLMs for generating further diagnostic tests; however, it neither benchmarks against physician performance nor analyzes discordant cases, so any systematic differences remain unknown^6^. Our qualitative analysis of the Capture criteria highlights how clinicians actually reason about diagnostic tests, which differs from simple diagnostic recall. Clinicians emphasized that identifying the DToR is rarely binary; it often involves proposing intermediate or pathway tests that narrow the differential before ordering the definitive study. To reflect this nuance, our framework distinguishes between direct matches (score = 1), where the DToR or its immediate precursor is named and partial pathway captures (score = 0.5), where a test strongly suggests the correct pathway but does not directly trigger the definitive diagnosis. Incorrect captures (score = 0) represent tests that are irrelevant or unrelated to the diagnostic trajectory.

This tiered scoring system acknowledges the collaborative, stepwise nature of diagnostic reasoning. For example, a chest X-ray is not definitive for lung cancer but was viewed by clinicians as a meaningful precursor that could appropriately lead to CT and biopsy; hence, it receives partial credit. In contrast, unrelated tests (e.g., ankle radiographs) provide no diagnostic value for the case at hand. Clinicians noted that this differentiation mirrors their own practice, where even non-definitive investigations can meaningfully guide the pathway toward the correct diagnosis, whereas irrelevant tests introduce noise and risk. This gradation better aligns evaluation with real-world clinical reasoning by rewarding steps that materially advance decision-making, not just end-point recall.

By conventional clinical-effect heuristics, OpenMedicine AI shows very large, practice-relevant gains. It rescued far more often than it gave up. At Capture@10 vs physicians, rescues outnumbered failure to rescues 99 vs 6 (matched OR 16.5), yielding approximately 31 fewer catastrophic misses per 100 cases and approximately 1 miss averted every 3.2 cases. Compared to AMIE, the ratio was 44 vs 6 (OR 7.33), corresponding to approximately 13 fewer catastrophic misses per 100 cases and approximately 1 every 8.0 cases. The safety margin appears already at Capture@3: 109 vs 15 against physicians (OR 7.27; approximately 31 fewer per 100; approximately 1 every 3.2 cases) and 61 vs 15 against AMIE (OR 4.07; approximately 15 fewer per 100; approximately 1 every 6.6 cases). Thus, OpenMedicine AI reduces catastrophic misses early and consistently, with larger gains when compared to physicians and smaller but reliable gains when compared to AMIE.

Differential lists are only one part of the diagnostic workflow. Diagnostic tests drive outcomes, cost, and safety. Concordance is necessary but not sufficient. Decision correctness and safety are what matter to patients and health systems. Our results suggest that gains typically pursued by either clinician-augmentation (time-heavy) or bespoke medical LLMs (cost-heavy) can be achieved through pipeline design. Here, we improved diagnostic actionability and DToR-aligned performance using general models with a deterministic, medicine-aware controller, requiring minimal clinician time for adjudication and no domain-specific pretraining. This offers a scalable, cost-efficient alternative for health systems. This capability may be valuable across time-sensitive settings, from emergency care to routine outpatient evaluation.

### Limitations

CPCs are designed as diagnostic conundrums and may not fully reflect real-world clinical workflows, which more often involve common conditions, iterative patient interactions, and contextual information absent from written case reports. Information is often missing, asynchronous, and conflicting. Moreover, CPCs frequently include details that would not be available at a new encounter, such as an emergency or urgent care visit. Without laboratory results or prior history, the information attainable in practice would be far more limited and unlikely to yield a concise, complete, and coherent narrative of the kind prepared at Massachusetts General Hospital. While OpenMedicine AI demonstrated strong performance under these enriched conditions, prospective validation in real-world settings will be essential to establish generalizability and clinical impact.

Evaluation of diagnostic test selection is similarly nontrivial. Although we used the OpenMedicine AI agent as an evaluator, reasonable disagreement may exist over whether a given test should be considered correct versus incorrect. Our Capture framework explicitly distinguishes between direct matches, partial pathway captures, and incorrect proposals. Clinicians naturally recognized these gradations when reviewing differentials, whereas baseline models tended to treat test selection as a binary outcome. By introducing partial credit for pathway-consistent tests (score = 0.5), our framework captures clinically meaningful reasoning steps that bridge from a broad differential to the definitive diagnostic test of record (score = 1). This provides a more nuanced and practice-aligned evaluation of diagnostic performance. Autonomous evaluation remains promising but unproven until we demonstrate, at minimum, that trends between human scoring and AI-based scoring remain consistent. Future work should explore alternative scoring schemes and assess their applicability across additional diagnostic modalities.

While the case reports included images and tables, OpenMedicine AI, like AMIE, had access only to the main text and still outperformed both physicians and AMIE. Future studies that more closely approximate real-world clinical scenarios, where information is incomplete and often fragmented across notes, labs, and prior visits, will need to consider how multimodal inputs such as imaging, laboratory data, and longitudinal clinical history can further inform model performance. CPC challenges alone are not a direct benchmark for ordering the minimum appropriate tests. Although our findings suggest that this capability is within reach and represents a logical next step, rigorous prospective studies in blinded clinical settings will be required to determine how test recommendations can most effectively support patient care.

## Conclusion

Generating a differential diagnosis is a critical first step in clinical case management, while the subsequent ordering of diagnostic tests carries direct implications for costs and patient outcomes. Our study shows that OpenMedicine AI can support both critical stages, not only producing accurate and comprehensive DDx lists but also guiding toward the most appropriate diagnostic tests. These findings indicate that a resource-efficient, model-agnostic diagnostic pipeline on frontier LLMs can exceed AMIE on both list accuracy and downstream diagnostic action, despite requiring far less model-specific development.

In sum, our third path, engineering the orchestration layer rather than the base model, delivers clinically meaningful improvements with materially lower time and cost than clinician-augmentation or bespoke model training. Prospective, multi-site studies should test whether higher Capture shortens time-to-DToR and reduces low-value testing. Although CPCs are valuable as rigorous benchmarks, they do not fully reflect real-world clinical workflows, which involve more common conditions, iterative patient interactions, and contextual cues absent from written cases. Prospective validation in real-world settings will therefore be essential to establish generalizability, safety, and clinical impact.

## Methods

### Case records

The case records of the Massachusetts General Hospital (MGH) are lightly edited transcriptions of the CPCs (Boston, MA) and are regularly published in the New England Journal of Medicine as diagnostic puzzles culminating in a definitive, pathology-confirmed diagnosis. For each CPC, OpenMedicine AI was prompted to generate a differential diagnosis (DDx) based solely on the history of present illness (HPI), excluding titles, solution text, discussion, figures, tables, references, or editorial content and any results from diagnostic tests. We leveraged these case reports to evaluate OpenMedicine AI’s ability both to generate accurate DDx lists and to identify diagnostic pathways through Capture@K, defined as the frequency with which an agent’s Top-n differentials led to the diagnostic test of record (DToR) or its immediate precursor. For this latter task, we developed an agent tool that mapped each proposed diagnosis to the diagnostic test it would trigger.

A total of 302 cases were selected, consistent with those used in prior benchmarking work. These cases spanned multiple specialties, with the largest representation from internal medicine (n = 159), followed by neurology (n = 42), pediatrics (n = 33), and psychiatry (n = 10). The HPI text was manually extracted from each report and used as input to OpenMedicine AI. To allow a head-to-head comparison with AMIE, which is a text-only model, accompanying images and tabular laboratory data from the CPCs were withheld from both models.

### OpenMedicine AI

OpenMedicine AI is a constraint-driven diagnostic agent and runs a deterministic six-step pipeline that mirrors expert reasoning while enforcing consistency and safety. The system structures cases, proposes mechanism-based hypotheses, and re-prioritizes them using evidence discriminators (organ distribution, imaging signatures, histology cues, lab anchors). External retrieval when used informs ranking but does not import answers. Outputs are standardized (final hypothesis, ranked differential, next-step plan) to fit clinical review and audit. Any development-time exposure predated this experiment and models were frozen prior to evaluation.

We evaluated OpenMedicine AI on each of the 302 NEJM case studies with the following prompt: “You are OpenMedicine AI, an AI language model invited to discuss clinicopathological cases with expert human clinicians. These cases come from the New England Journal of Medicine (NEJM). Your goal is to generate the most accurate and specific diagnosis possible based only on the case description. Your output will be compared against human expert performance and pathologic confirmation.”

## RULES & FORMAT

Task

After reading the case, you must provide:

A. Most Likely Diagnosis (specific and final)
B. Ranked Differential Diagnosis (as specific as possible; ordered by likelihood)

You do not need to explain your reasoning, just list diagnoses.

Algorithmic Procedure

Extract Structured Case Features (No diagnostic labels in this step; descriptive only.)

Age / Sex

Chief Complaint

Time Course (acute, subacute, chronic)

Associated Symptoms / Findings

Vitals & Labs (salient abnormalities only)

Imaging (key patterns and distributions)

Biopsy / Pathology (descriptive, if present)

Treatments Tried / Responses Missing / Negative Features (explicitly absent but decision-relevant)

Immunologic / Serologic Markers (values/positivity only)

Diagnosis-oriented Query OpenMedicine AI distills the features into a single question: “What diagnosis best explains [key findings] and [course/response]?”

Decision Rules (applied before final output)

Syndrome → Underlying disease. If a candidate is a syndrome (e.g., Sweet’s, paraneoplastic pemphigus), replace or supplement with the most likely root cause (e.g., AML, NHL, Castleman disease). Global fit. The lead diagnosis must account for systemic and imaging findings; otherwise re-rank. Mimic cleanup. Down-weight common mimics (e.g., SLE) when inconsistent with the feature set. Therapy signal. Incorporate treatment response/non-response (e.g., steroid-responsive infiltrative processes).

Cross-system consistency. Ensure the Top diagnosis explains multi-organ patterns. Heuristic lens used for ranking: “Begin with the key clinical finding (e.g., thrombocyTopenia, abdominal pain, transaminitis, weakness). Classify mechanisms: destruction, obstruction, infiltration, suppression, malfunction, deficiency, disconnection and consider etiologies across congenital, infectious, autoimmune, neoplastic, toxic/metabolic, and degenerative domains.”

Reproducibility. Model/version and parameters are fixed at study start; per-case inputs and outputs are logged. No explanatory text is produced at inference time.

Output Specification (per case)

Most Likely Diagnosis One line, specific, final entity (e.g., “IgG4-related disease, systemic”).

Ranked Differential Diagnoses — Google Sheets–ready single column (CSV header, no numbering)

Diagnosis

IgG4-related disease

Sarcoidosis

Idiopathic orbital inflammation (non-IgG4)

Orbital lymphoma (MALT or DLBCL)

Granulomatosis with polyangiitis

Castleman disease

Tuberculous orbital mass

Use one column titled Diagnosis. Avoid punctuation within entries; each diagnosis is a standalone row.

Case Reporting Template

Case Features (extracted):

Age / Sex:

Chief Complaint:

Time Course:

Associated Symptoms / Findings:

Vitals & Labs (salient):

Imaging (salient):

Biopsy / Pathology (descriptive):

Treatments Tried / Responses:

Missing / Negative Features:

Immunologic / Serologic Markers:

Most Likely Diagnosis: [Final specific diagnosis]

### Automated evaluation

For each CPC case, we generated a ranked list of N differential diagnoses (DDx). We then used an adjudicator (GPT-4o) to determine (i) whether the model’s predicted diagnosis exactly matched the ground-truth diagnosis and (ii) which item(s), if any, in the DDx list matched the ground truth using the following prompt:

1. Check whether the predicted differential diagnosis exactly matches the true diagnosis
2. Identify which item(s) in a provided differential-diagnosis (DDx) list match the true diagnosis. The lowest number will be recorded to represent Top-n for that CPC.

### Capture@K criteria

To assess whether model-generated differentials (DDx) guided the case toward the definitive diagnostic pathway, we defined a Capture@K framework. For each case we identified the diagnostic test of record (DToR), the single most definitive test establishing the ground-truth diagnosis, or its immediate precursor when a precursor test is standard (e.g., chest CT preceding bronchoscopic biopsy for lung cancer).

Each DDx item was mapped to the test it would trigger, and a Capture score was assigned using a three-tier scale: 1 if the DToR or its immediate precursor was captured; 0.5 if the proposed test strongly supported the correct pathway but was not the immediate precursor; 0 if unrelated to the DToR. Capture was evaluated at K∈{3, 5, 10} (Capture@3/5/10), corresponding to whether the DToR (or precursor) appeared within the Top-n differentials. For each case, Capture@K was the highest score achieved among the Top-n items.

### Statistical analysis

For Top-1 and Top-10 diagnostic-list accuracy on paired cases, differences between OpenMedicine AI, AMIE and physicians were tested via two-sided McNemar’s exact test.

For the win-or-tie frequency analysis, we report a frequency-based Capture@K (“win-or-tie”, ≥): the proportion of cases for which one model’s Capture@K matched or exceeded a comparator, with Wilson 95% CIs.

Because Capture scores are ordinal {0, 0.5, 1}, the Wilcoxon signed-rank test (paired; two-sided, α=0.05) was the primary test comparing models; we report Wilcoxon V and P. Paired superiority was tested using McNemar’s test (exact mid-P when discordant counts were small).

For discordant pairs, we report the matched odds ratio (rescues/failure to rescues) with a 95% CI. Primary paired comparisons were prespecified at Capture@3, Capture@5 and Capture@10 under the strict scoring rule (success = 1.0; 0.5 and 0 counted as miss). We defined a rescue when OpenMedicine AI was correct and the comparator was not, and a failure to rescue for the reverse; ties were excluded. We report the matched odds ratio (OR) = rescue/failure to rescue with 95% CIs from the log-OR (Wald) formula exponentiated to the OR scale. P values are from two-sided McNemar’s exact test on discordant pairs.

A catastrophic miss was counted when the comparator scored 0 and OpenMedicine scored 1 (strict). We report the reduction per 100 cases with Wilson 95% CIs for the underlying proportion (wins / 302), and compute NNA = 100 / (reduction per 100). For interpretability, we also determined the paired risk difference (RD) in strict success rates; 95% CIs for RD were obtained by paired bootstrap over cases.

## Data Availability

All data produced in the present study are available upon reasonable request to the authors.

## Data availability

The case reports in this study were licensed from the New England Journal of Medicine. Because they are copyrighted, we cannot share the texts ourselves. Readers may obtain the original case materials directly from the journal.

## Supplementary information

**Table S1:**
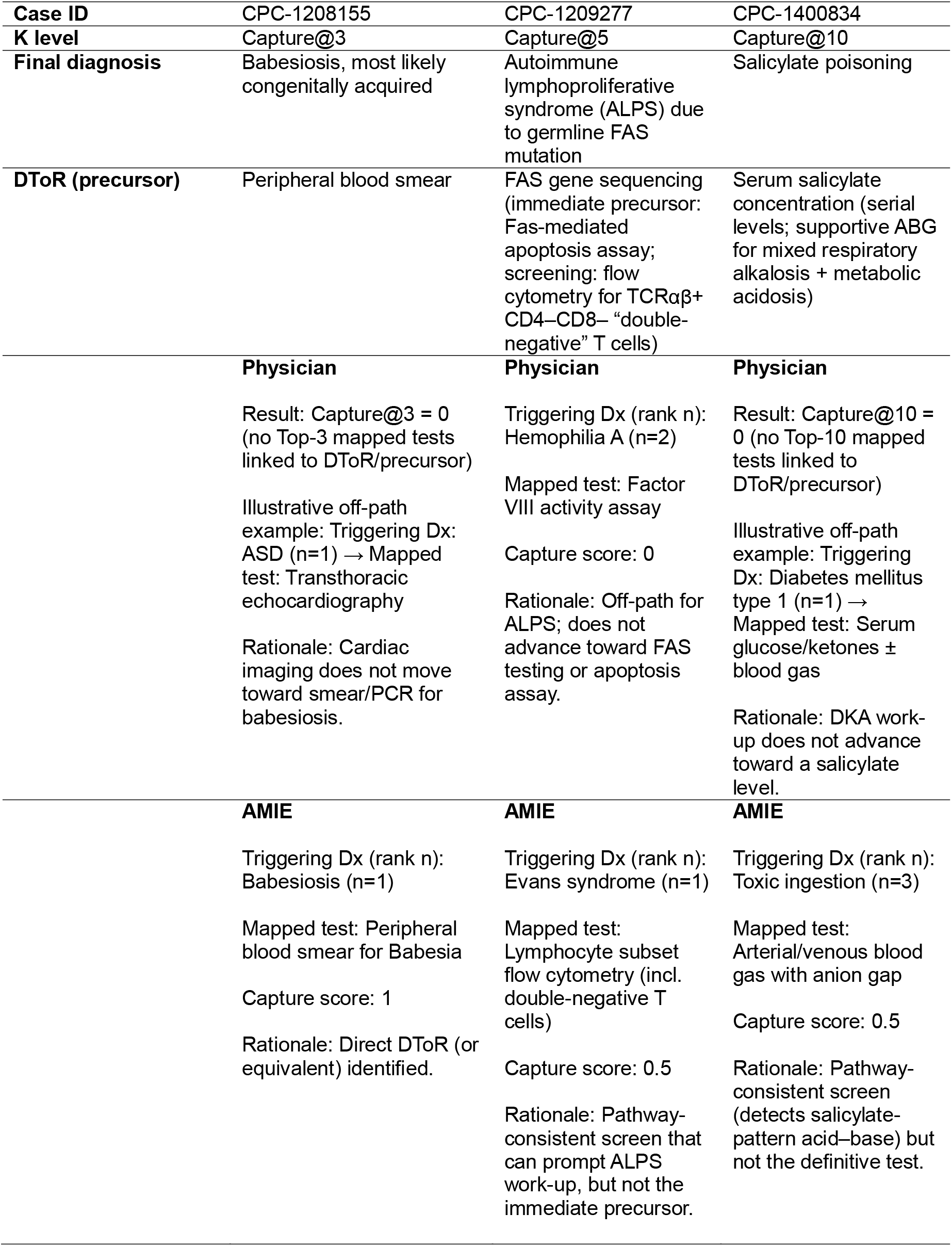
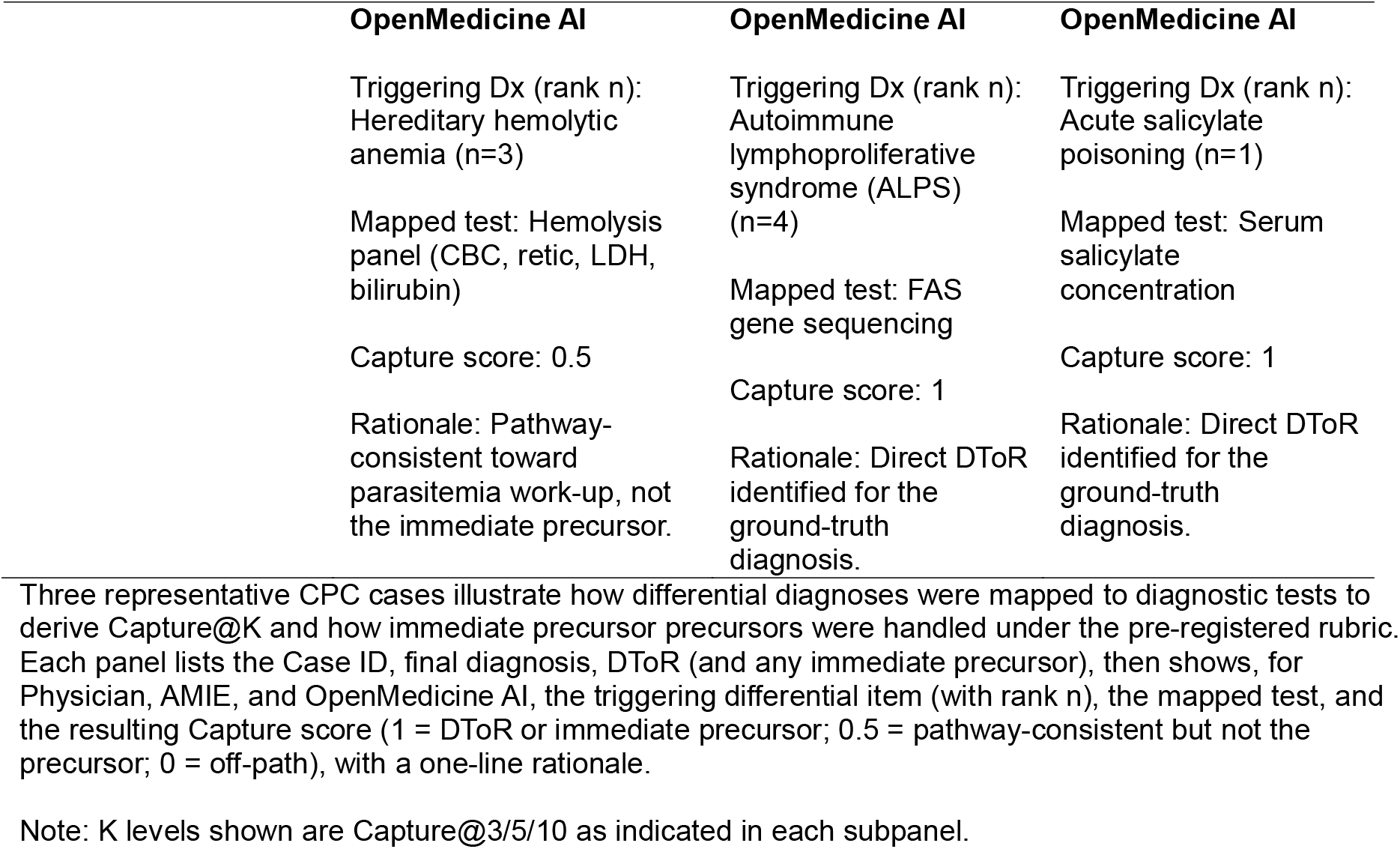
Worked examples of differential-to-test mapping used to compute Capture@K.

## References

1 Goh, E. et al. GPT-4 assistance for improvement of physician performance on patient care tasks: a randomized controlled trial. Nat Med 31, 1233–1238, doi:10.1038/s41591-024-03456-y (2025).

2 Goh, E. et al. Large Language Model Influence on Diagnostic Reasoning: A Randomized Clinical Trial. JAMA Netw Open 7, e2440969, doi:10.1001/jamanetworkopen.2024.40969 (2024).

3 Tu, T. et al. Towards conversational diagnostic artificial intelligence. Nature 642, 442–450, doi:10.1038/s41586-025-08866-7 (2025).

4 Singhal, K. et al. Toward expert-level medical question answering with large language models. Nat Med 31, 943–950, doi:10.1038/s41591-024-03423-7 (2025).

5 Hayat, H. et al. Toward the Autonomous AI Doctor: Quantitative Benchmarking of an Autonomous Agentic AI Versus Board-Certified Clinicians in a Real World Setting. medRxiv, 2025.2007.2014.25331406, doi:10.1101/2025.07.14.25331406 (2025).

6 Chen, X. et al. Enhancing diagnostic capability with multi-agents conversational large language models. NPJ Digit Med 8, 159, doi:10.1038/s41746-025-01550-0 (2025).

7 Kanjee, Z., Crowe, B. & Rodman, A. Accuracy of a Generative Artificial Intelligence Model in a Complex Diagnostic Challenge. JAMA 330, 78–80, doi:10.1001/jama.2023.8288 (2023).

